# The role of peripheral immunity in ALS: a population-based study

**DOI:** 10.1101/2022.09.06.22279493

**Authors:** Maurizio Grassano, Umberto Manera, Fabiola De Marchi, Paolo Cugnasco, Enrico Matteoni, Margherita Daviddi, Luca Solero, Alessandro Bombaci, Francesca Palumbo, Rosario Vasta, Antonio Canosa, Paolina Salamone, Giuseppe Fuda, Federico Casale, Letizia Mazzini, Andrea Calvo, Cristina Moglia, Adriano Chiò

## Abstract

**Background:** Systemic inflammation has been proposed as a relevant mechanism in Amyotrophic Lateral Sclerosis (ALS). Recent evidence shows that an increased inflammatory status correlates with survival in ALS. Still, comprehensive data on ALS patients’ innate and adaptive immune responses and their effect on the clinical phenotype are lacking. Here, we investigate the role and characteristics of systemic immunity in a population-based ALS cohort using readily available hematological indexes that reflect changes in innate and adaptive immunity.

**Methods:** We collected the complete blood count (CBC) at diagnosis in ALS patients from the Piemonte and Valle d’Aosta Register for ALS (PARALS) from 2007 to 2019. Demographic and clinical data were collected using registry data. Leukocytes populations, neutrophil-to-lymphocyte ratio (NLR), platelet-to-lymphocyte ratio (PLR), systemic immune-inflammation index (SII) and lymphocyte-to-monocyte ratio (LMR) were derived from CBC. All variables were analyzed for association with clinical features in the entire cohort and then in sex- and age-based subgroups. Logistic, linear and Cox regression models were used as appropriate.

**Results:** After adjustment for relevant covariates, neutrophils (p=0.001) and markers of increased innate immunity (NLR, p=0.008 and SII, p=0.006) were associated with a faster disease progression. Similarly, elevated innate immunity markers correlated with worse pulmonary function and shorter survival. Sex-based differences emerged, as the prognosis in women also correlated with a low lymphocyte (p=0.045) and a decreased LMR (p=0.013). ALS patients with cognitive impairment exhibited lower levels of monocytes (p=0.0415) and, although only in later-onset ALS (age at onset > 70 years), lower lymphocytes (p=0.006) and increased NLR (p=0.021) and SII (p=0.030).

**Conclusions and Relevance:** Our results confirm that a dysregulated systemic immune system participates in ALS progression. More specifically, an elevated innate immune response is associated with faster progression and reduced survival. The immune response varied according to sex and age, thus prompting the speculation that involved immune pathways are patient-specific. Finally, we observed that ALS patients with greater cognitive impairment showed a reduction in monocytes count. Those data revealed that systemic inflammation plays a multifaceted role in ALS: further studies will help translate those findings into clinical practice or targeted treatments.

## INTRODUCTION

Neuroinflammation has been postulated to be a relevant mechanism in the neurodegenerative process in Amyotrophic Lateral Sclerosis (ALS)^1^. Previous studies found that the frequency and activation of immune cell populations such as T cells, monocytes and neutrophils have been associated with disease severity and rate of disease progression^2–5^. Innate immunity involves various types of cells of the myeloid lineage, including dendritic cells, monocytes, macrophages, polymorphonuclear cells, mast cells, innate lymphoid cells, and natural killer (NK) cells^6,7^. Adaptive immunity is characterized by two types of lymphocytes, T and B cells^6^. The most easily and generalizable method to qualify peripheral immunity is blood cell counting of differential leukocytes. Furthermore, derived ratios including neutrophil-to-lymphocyte ratio (NLR), platelet-to-lymphocyte ratio (PLR), systemic immune-inflammation index (SII) and lymphocyte-to-monocyte ratio (LMR), were thought to even better reflect the strength of peripheral immunity^8^. There have been studies suggesting that these ratios could serve as systemic biomarkers of ALS prognosis^9–11^. Because systemic inflammation is a complex and a multifactorial process, however, a better interpretation of the immune profile in ALS patients requires the simultaneous study of different biomarkers.

Here, we describe the systemic immune role in ALS patients by analyzing complete blood count (CBC) - derived parameters in a large population-based cohort. A better understanding of the systemic immune response in the context of ALS and frontotemporal dementia (FTD) is not only important to unravel disease mechanism, but potentially to serve as biomarker of disease activity.

## MATERIALS AND METHODS

### Study population

All patients with ALS in the Piemonte and Valle d’Aosta regions of Italy (n = 1784), identified through the Piemonte and Valle d’Aosta Register for ALS and diagnosed between January 1, 2007, and December 31, 2019, were eligible for enrollment in the study. All patients met the revised El Escorial diagnostic criteria for definite and probable laboratory-supported ALS. A complete clinical history, including previous disease (especially history of cancer and inflammatory disease) and smoking habit (current, past or never smoker), was collected for each patient. We restricted analyses to available blood count data within 3 months of ALS diagnosis. In addition, participants with morbidities that could influence leukocyte differential counts, including malignant neoplasms, disease of the blood and blood-forming organs, autoimmune disease, and chronic inflammatory diseases were excluded.

### Peripheral immunity

We extracted baseline count data of neutrophils, monocytes, platelets, and lymphocytes. Further we calculated four ratios based on peripheral blood cell counts including NLR (neutrophils/ lymphocytes), PLR (platelets/lymphocytes), SII (neutrophils*platelets/lymphocytes), and LMR (lymphocytes/monocytes). The increased level of the neutrophils, monocytes, NLR, PLR, and SII reflect the relatively higher peripheral innate immunity, while the increased level of the lymphocytes and LMR reflect the relatively higher peripheral adaptive immunity.

### Clinical features

Disease severity was assessed with the Amyotrophic Lateral Sclerosis Functional Rating Scale– Revised scale (ALSFRS-R). In addition, the decline rate for ALSFRS-R score and its 4 sub-scores (bulbar, fine motor, gross motor, and respiratory) was calculated as the monthly number of points lost from symptom onset to the time of diagnosis. Pulmonary function tests, including forced vital capacity (FVC), were performed at diagnosis. Respiratory dysfunction was defined as FVC < 75% of the predicted value. Body mass index (BMI) was calculated as weight in kilograms divided by height in meters squared, and its decline was calculated as monthly the difference in premorbid BMI and BMI at diagnosis, divided by months from symptom onset to diagnosis. Patients’ cognitive status was classified as previously described according to the revised ALS-FTD Consensus Criteria into five categories: ALS with normal cognition (ALS-CN); ALS with behavioral impairment (ALS-Bi); ALS with cognitive impairment (ALS-Ci); ALS with cognitive and behavioral impairment (ALS-CBi); ALS with FTD (ALS-FTD).

Survival was calculated from onset to death or censoring date (December 31st, 2019) using the Kaplan-Meier method. Patients with tracheostomy were coded as deceased on the date of the procedure.

### Statistical analyses

Baseline characteristics of participants were analyzed as mean and standard deviation (SD) for continuous variables and as numbers and percentages for categorical variables. Given the high interpersonal variability of baseline FVC values, we elected to assess respiratory function as a dichotomous variable (FVC < 75% of predicted value). Similarly, ALSFRS-R progression rate, Prior to statistical analysis, peripheral immunity cell counts were log-transformed and standardized to Z scores for comparison of effect sizes between exposures (Z = (value − mean)/SD) such that the hazard ratio (HR) represents the predicted effect of per SD increment of the peripheral immunity markers. In the primary analysis, we performed the models unadjusted, then adjusted for age, sex, smoking history, presence of bulbar dysfunction (score < 4 in either item 1, 2 or 3 of the ALSFRS-R scale) and body mass index (BMI). Association between peripheral immunity markers and survival were examined with multivariable Cox proportional hazard regression models. The P values were further adjusted to control the false discovery rate at 5% using the Benjamini –Hochberg procedure (labeled as Q values). For each biomarker, we also performed the analysis excluding patients with extreme values (>mean ± 2SD) in at least one of the other exposures, thus correcting for potential correlation across different biomarkers. In stratified analyses, we estimated differences for each exposure across different age groups (<65 years, 65 – 75 years, >75 years) and sex.

### Ethical approval

The study design was approved by the institutional ethical committees of Azienda Ospedale Università, Città della Salute e della Scienza, and Azienda Ospedale Università Maggiore di Novara. All patients provided written informed consent.

## RESULTS

### Population characteristics

Among the 1784 patients included in the PARALS Registry in the study period, 1452 (81.5%) subjects were eligible for analysis. The final cohort consisted of 652 (44.9%) females and 799 (55.1%) males. Median age at disease onset was 69.5 years (IQR 61.4-75.7); bulbar onset was observed in 489 (33.7%) cases. Descriptive statistics of the participants are summarized in eTable 1. Patients excluded from the study did not differ in term of demographic or other clinically relevant variables (eTable 1). Blood-count derived data are reported in eTable 2.

### Baseline

After adjustment for age, sex, presence of bulbar involvement, smoking habits, and BMI, per SD increment of neutrophils, the main components of innate immunity, was associated with higher rates of disease progression (β 1.03, 97.5% CI 1.01-1.05, P = 0.001) (eTable 4 and Figure 1B). Similar findings were observed in higher ratios of the markers reflecting innate immunity over adaptive immunity, including NLR (per SD increment β1.03, 97.5% CI 1.01-1.05, P = 0.008) and SII (per SD increment β 1.03, 97.5% CI 1.01-1.05, P = 0.006). No significant effect was found for platelets and PLR on progression rates. The level of lymphocytes, the core adaptive immunity components, was not associated with progression rates. Results were not significant also for LMR. Patients with a faster disease progression also exhibited an increase in peripheral monocytes (β 1.05, 97.5% CI 1-1.09, P = 0.0410). Importantly, no systemic inflammation signature was affected by bulbar dysfunction (eTable 3B).

**Figure 1.**
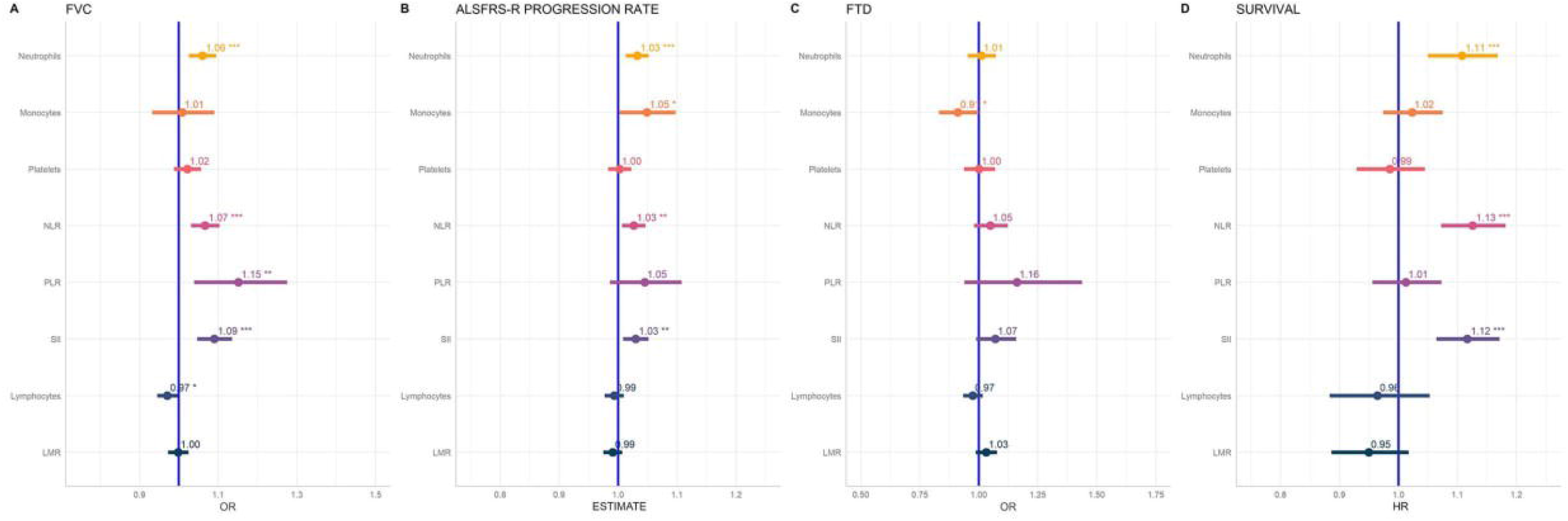
Overall association of peripheral immune markers with FVC < 75% of predicted value (A), ALSFRS-R progression rate (B), cognitive impairment (C) and survival (D). OR odds ratio, HR hazard ratio, 97.5% CI 97.55% confidence interval, NLR neutrophil-to-lymphocyte ratio, PLR platelet-to-lymphocyte ratio, SII systemic inflammatory index, LMR lymphocyte-to-monocyte ratio

As for pulmonary function, a consistent association with respiratory failure (here defined as FVC% < 75% of the predicted value) was found in innate immunity, as presented for neutrophils (per SD increment OR 1.06, 97.5% CI 1.03 -1.09, P =0.000473) NLR (per SD increment OR 1.07, 97.5% CI 1.03 -1.10, P = 0.000181), PLR (per SD increment OR 1.15, 97.5% CI 1.04 - 1.28, P=0.00712) and SII (per SD increment OR 1.09, 97.5% CI 1.05 -1.14, P =0.0000351), respectively (Figure 1A). Conversely, an association of adaptive immunity with higher FVC values was observed only in unadjusted models for lower lymphocyte levels (OR 0.97, 97.5% CI 0.95-0.99, P = 0.02965).

Unsurprisingly, given the association with disease progression rates and respiratory function, innate immunity markers also correlated with a shorter disease survival (Figure 1D). No direct association was found regarding altered adaptive/regulatory pathways in ALS subjects with a shorter survival.

Concerning FTD and cognitive symptoms incidence, lower levels of monocytes were related to increased risk of FTD (per SD increment OR 0.92, 97.5% CI 0.86-0.99, P 0.0474) (eTable 4) and cognitive impairment (per SD increment OR 0.84, 97.5% CI 0.73- 0.97, P 0.01704) (Figure 1C).

### Peripheral immunity and ALS phenotypes across different ages and sex

To understand whether the observed relationship between peripheral immunity and ALS clinical features was influenced by age or sex, both known modifiers of the immune response, we performed a series of subgroup analyses.

Firstly, we set apart the population by sex (eTable 5). The effect of innate immunity was explicit for disease progression and survival either in males or females (Figure 2D). Significant differences between the two sexes emerged regarding the mediator of immunity, as evidence of a detrimental effect of low lymphocyte count on prognosis was found in females (per SD increment Cox-HR 0.84, 97.5% CI 0.70 – 0.99, P=0.04503) but not in males. Similar results were obtained for LMR (per SD increment Cox-HR 0.84, 97.5% CI 0.72 – 0.96, P=0.0129). By contrast, only innate immunity marker that accounts for an increased neutrophil count were correlated with disease progression and prognosis in males. Low monocytes were also proven to slightly increased the incidence of cognitive deficits only in females (per SD increment OR 0.79, 97.5% CI 0.63 - 0.99, P=0.04613) (Figure 2C). Curiously, the opposite trend was observed in progression rate, where we observed a more severe disease progression when monocyte levels increased (per SD increment β 1.09, 97.5% CI 1.02-1.16, P=0.00948) (Figure 2B).

**Figure 2.**
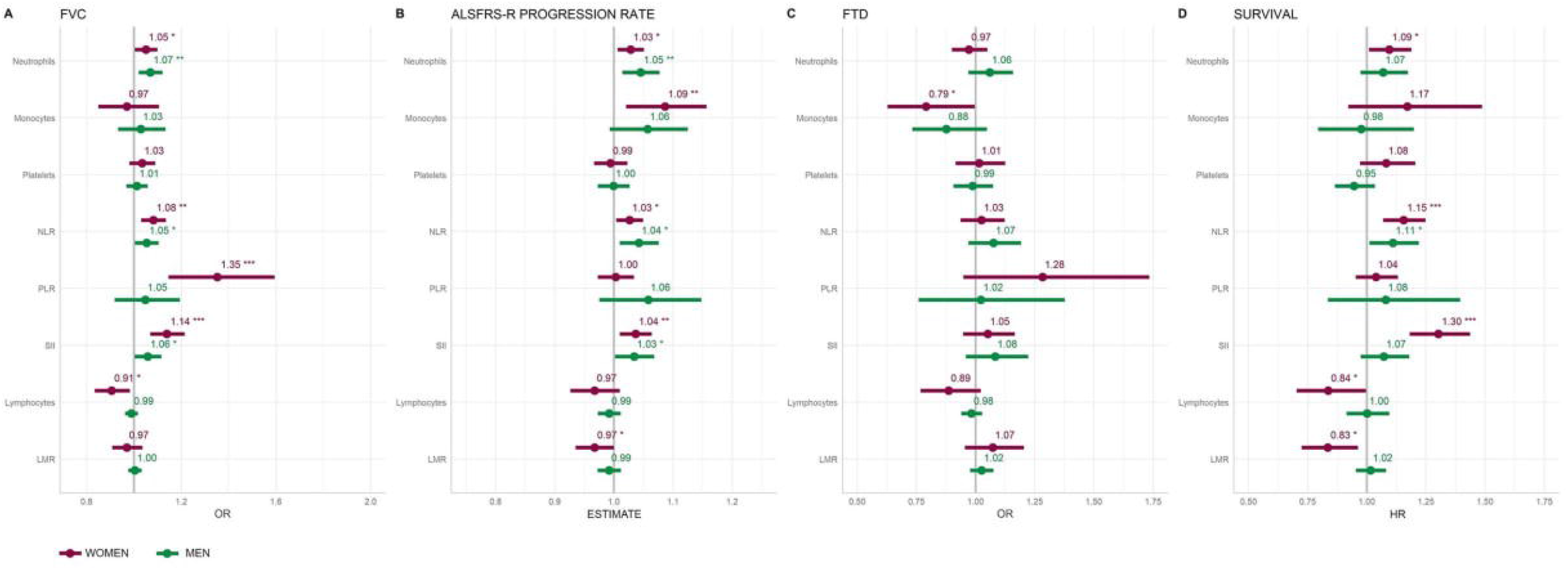
Sex-stratified association of peripheral immune markers with FVC < 75% of predicted value (A), ALSFRS-R progression rate (B), cognitive impairment (C) and survival (D). OR odds ratio, HR hazard ratio, 97.5% CI 97.55% confidence interval, NLR neutrophil-to-lymphocyte ratio, PLR platelet-to-lymphocyte ratio, SII systemic inflammatory index, LMR lymphocyte-to-monocyte ratio

Subsequently, we divided the included participants into three groups by age (<60 years, 60-70 years, >70 years). In participants younger than 60 years (Figure 3, eTable 6), only one innate immunity marker (the SII) increased with worsening respiratory function, but not with disease progression or survival (FIG 3A, 3B and 3D). Low-monocytes (OR 0.79, 97.5% CI 0.63 - 0.99, P=0.04090) and therefore an increased LMR (OR 1.11, 97.5% CI 1.03 – 1.19, P=0.00616) were found to be associated with cognitive impairment in young-onset ALS cases. Surprisingly, lymphocytes (OR 0.70, 97.5% CI 0.54 0.90, P=0.00574) are decreased in older patients (> 70 years) with cognitive impairment (Figure 3C). In addition to reduced lymphocyte, we also observed increased NLR (OR 1.15, 97.5 CI % 1.02-1.30, p=0.021) and SII (OR 1.13, 97.5 CI % 1.01-1.26, p=0.030) in older ALS patients with an overlapping FTD (eTable 6). When it moves to 60–70 years and over 70 years, significant detrimental roles were found for the increase of all innate immunity markers accounting for neutrophil levels (neutrophils, NLR and SII) in disease progression and survival (Figure 3). Those results suggest that the effect of innate immunity is less pronounced in younger patients.

**Figure 3.**
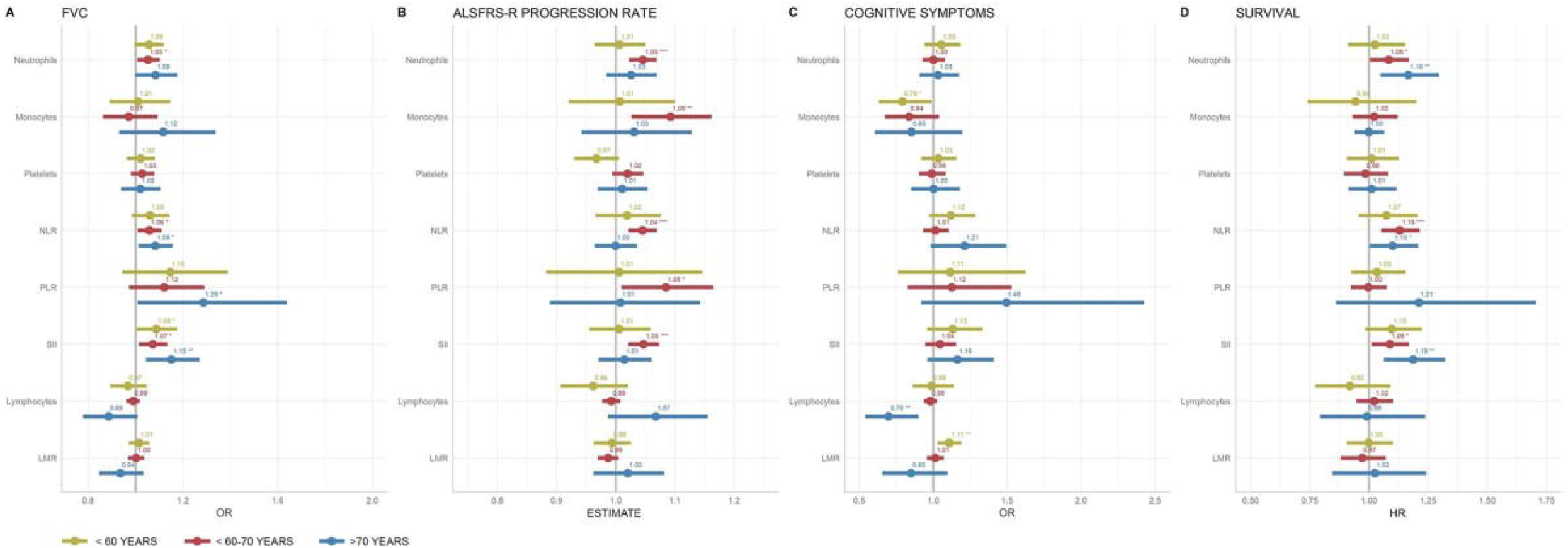
Age-stratified association of peripheral immune markers with FVC < 75% of predicted value (A), ALSFRS-R progression rate (B), cognitive impairment (C) and survival (D). OR odds ratio, HR hazard ratio, 97.5% CI 97.55% confidence interval, NLR neutrophil-to-lymphocyte ratio, PLR platelet-to-lymphocyte ratio, SII systemic inflammatory index, LMR lymphocyte-to-monocyte ratio

## Discussion

To our knowledge, this is the first study to comprehensively evaluated multiple CBC-derived inflammatory indexes, including novel candidate markers, in a large population-based ALS cohort. A graphic summary of our results can be found in Figure 4.

**Figure 4.**
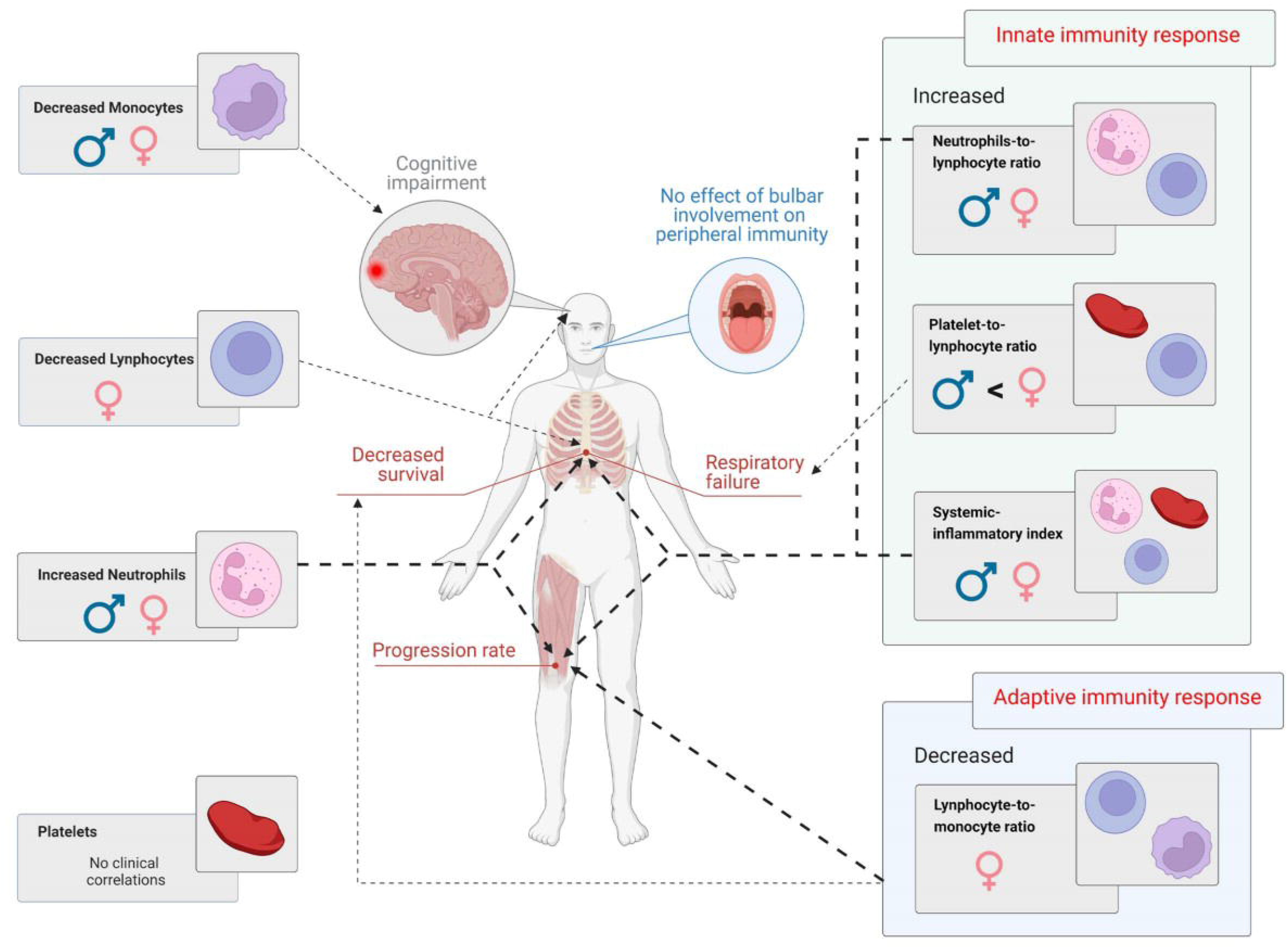
Graphical summary of study results.

We found that elevated innate immune inflammatory status, reflected by increasing peripheral neutrophils and a higher ratio of neutrophils to lymphocytes, is associated with reduced survival and faster disease progression. These data are consistent with previous studies that have linked neutrophil levels to disease progression and NLR with a higher mortality^5,9,10^. The associations of inflammatory response on disease progression, pulmonary function and prognosis are independent of the presence of smoking habit or bulbar dysfunction at diagnosis. In this regard, it is noteworthy that higher levels of all innate immune response markers are not associated with involvement of bulbar function, because it excludes potential confounding factors such as silent infection (i.e. pneumonia resulting from silent aspiration) that are common complications in patients with bulbar symptoms and which may trigger the inflammatory response. We also observed that other inflammatory biomarkers, namely decreased absolute lymphocytes count, higher monocytes count and their increased ratio (lymphocyte-to-monocyte), are linked to disease severity and worse prognosis in ALS patients, especially women. This correlation between a reduction of total lymphocytes and a shorter survival showed that the adaptive immune response is also a key player in the systemic response in ALS cases: unfortunately, given the nature of our study, we are unable to detect any changes in lymphocytes subpopulations that have been extensively described inALS^2,3,5,12–15^.

Our overall and stratified data demonstrated that an overregulated innate immune system at diagnosis is associated with a more aggressive disease course and confirms that neutrophils are protagonists of inflammatory response in ALS across all age groups and sex. However, according to our results, peripheral immune involvement in ALS is multifaceted and different mechanisms and leukocytes population are involved. Consequently, a single marker is not sufficient to capture the overall peripheral immune dysregulation and inflammatory status in ALS, and the evaluation of multiple markers is necessary to disentangle the complex underlying mechanism involved in ALS pathogenesis and progression.

Another novel finding from our study was the inverse association between monocyte levels and the degree of cognitive impairment in patients with ALS. Patients with greater cognitive impairment showed a reduction in all monocyte subpopulations. Our study could not unravel whether these changes are contributing to the ALS-FTD spectrum or a result of it. However, there are several reports of increased CNS infiltration of monocytes in ALS patients, and prior studies have speculated that a reduction in peripheral blood monocytes could be due to the recruitment and infiltration of these cells into theCNS^3,16^. Moreover, deregulation of the different populations of peripheral blood monocytes and their subsequent entry into the CNS may contribute to the neurodegenerative process or have a modifying role in the context of ALS^17^.

In this context, it is worth noting that monocytes manifested an opposite trend regarding motor progression (the faster the functional decline, the higher the level of circulating monocytes): this finding questions whether we are observing two different phases of a single inflammatory response or rather the activation of different pathways. Although it remains unclear whether monocytes play a primary or secondary role in neurodegeneration, our results point to a specific immune response in the subset of ALS patients with overlapping cognitive impairment or FTD. While further evidence is necessary, our data confirm that monocytes may represent an attractive target to study disease-associated neuroinflammatory processes.

An effect on the risk of cognitive impairment was detected for adapted immunity: among older ALS patients (> 70 years at diagnosis) lymphocytes count was decreased; younger ALS cases (< 60 years at diagnosis) exhibited instead lower LMR levels, although this effect might be driven by the reduction in peripheral monocytes. Conversely, despite their correlation with motor progression in the entire cohort, innate immunity markers were associated with FTD-like features only in late-onset ALS patients. However, this correlation might reflect the changes in the lymphocyte populations.

Finally, our findings suggest that the mechanism of immune response in ALS varies according to sex. When the analysis was stratified by sex, we found that while the effect of neutrophil levels remained consistent, low peripheral lymphocytes were associated with respiratory dysfunction and shorter survival. The cause for this discrepancy is currently not clear, although sex-based differences in peripheral immunity in ALS have been reported before^18^. While further evidence is required to ponder whether sex-based immunological differences contribute to the variation in the clinical manifestation that characterizes female and male ALS subjects^19^, our results emphasized that sex is a variable that should be considered when studying phenotypes and biomarkers in motor neuron disorders.

Our study has some limitations. Since we converted values from cell counts and ratio to z-score, other efforts will be required to find clinically relevant and consistent cut-offs that could establish those indexes as reliable biomarkers in clinical practice. Furthermore, because of the retrospective nature of our cohort, the present study is not designed to answer whether targeting the peripheral immune system with therapeutic interventions could be beneficial for ALS patients.

Indeed, whether the observed alteration in systemic immunity is a cause or a consequence of disease progression and phenotypic variability in ALS is still unclear. For instance, neutrophils have several putative roles in ALS which could also differ according to the disease stage: most prominently, neutrophils may either have a pro-inflammatory role in driving neurodegeneration and promoting the breakdown of the brain–spinal cord barrier, or they could be involved in neuronal repair and increment in response to CNS damage.

Additionally, a dysregulated immune response may also indirectly contribute to increased mortality in ALS through its correlation with respiratory function: chronic inflammation is known to promote a decline in lung function, including forced vital capacity (FVC)^20^. In this regard, markers of inflammatory status like the neutrophil to lymphocyte ratio may reflect the adverse respiratory associations of exogenous exposures with lung function, like smoking and other nonsmoking exposures with respiratory outcomes (eg, air pollution)^21^.

While smoking habits did not significantly influence the inflammatory status in our cohort, it cannot be excluded that immune system functioning acts as a mediator of one of the several other environmental factors (nutrition status, pollutant exposures, physical activity, and the composition of microbiome among others) that are supposed to contribute to ALS.

This study validated the existence of peripheral immune abnormalities in ALS patients and explored the association between peripheral immunity and the clinical characteristics of ALS. We demonstrated that at diagnosis ALS patients with an increased innate immune activity had a faster progression of motor and cognitive symptoms and reduced survival times. Different peripheral immune cells may specifically contribute to the multifaceted mechanisms and clinical features in ALS, as observed with monocytes and cognitive deficits. Finally, immune response in ALS may be sex-specific, as adaptive immune response seems to play a more prominent role in females rather than in male ALS subjects. In conclusion, ALS is a disorder with extensive systemic pro-inflammatory responses: further studies are required to translate our findings into novel clinical markers and potential therapeutic targets.

## Supporting information

eTables

## Data Availability

Data are available upon reasonable request to the corresponding author by interested researchers.

## Fundings

This work was supported by the Italian Ministry of Health (Ministero della Salute, Ricerca Sanitaria Finalizzata, grant RF-2016-02362405); the Progetti di Rilevante Interesse Nazionale programme of the Ministry of Education, University and Research (grant 2017SNW5MB); the European Commission’s Health Seventh Framework Programme (FP7/2007–2013 under grant agreement 259867); and the Joint Programme–Neurodegenerative Disease Research (Strength, ALS-Care and Brain-Mend projects), granted by Italian Ministry of Education, University and Research. This study was performed under the Department of Excellence grant of the Italian Ministry of Education, University and Research to the “Rita Levi Montalcini” Department of Neuroscience, University of Torino, Italy, and to the Department of Health Sciences, University of Eastern Piedmont, Novara, Italy. The funders had no role in data collection or analysis and did not participate in writing or approving the manuscript

## Data availability statement

Data are available for interested researchers upon reasonable request to the corresponding author.

## Conflict of Interest Disclosures

Maurizio Grassano, Umberto Manera, Fabiola De Marchi, Paolo Cugnasco, Enrico Matteoni, Margherita Daviddi, Luca Solero, Alessandro Bombaci, Francesca Palumbo, Rosario Vasta, Antonio Canosa, Paolina Salamone, Giuseppe Fuda, Federico Casale, Letizia Mazzini, Cristina Moglia: no disclosures.

Andrea Calvo has received a research grant from Cytokinetics.

Adriano Chiò serves on scientific advisory boards for Mitsubishi Tanabe, Roche, Denali Pharma, Cytokinetics, and Amylyx.

## Notes

### Funding Statement

Italian Ministry of Health (Ministero della Salute, Ricerca Sanitaria Finalizzata, grant RF-2016- 02362405); Ministry of Education, University and Research (Progetti di Rilevante Interesse Nazionale programme, grant 2017SNW5MB); European Commissions Health Seventh Framework Programme (FP7/20072013 under grant agreement 259867).

### Author Declarations

The study design was approved by the institutional ethical committees of Azienda Ospedale Universita, Citta della Salute e della Scienza, and Azienda Ospedale Universita Maggiore di Novara. All patients provided written informed consent.

