## Supplementary material for "The role of peripheral immunity in ALS: a population-based study": eTables

**eTABLE 1. Descriptive statistics of the cohort**

|  | **All patients (n=1784)** | **Patient included in the study (n=1451)** |  |
| --- | --- | --- | --- |
|  | ***Median (IQR)*** | ***Median (IQR)*** | ***Mann-Whitney p-value*** |
| **Age at diagnosis** (years) | 69.8 (61.8-76.0) | 69.5 (61.4-75.7) | 0.418 |
| **Weight loss** (onset-diagnosis, kilograms) | 3.0 (0.0-8.0) | 3.0 (0.0-7.0) | 0.499 |
| **FVC** (%) | 88.0 (68.0-103.0) | 89.0 (70.0-103.0) | 0.322 |
| **ALSFRS-R RATE** (points lost/month) | 0.69 (0.33-1.43) | 0.66 (0.33-1.33) | 0.256 |
| **Overall survival** (months) | 18.2 (8.2-33.4) | 19.1 (9.0-35.1) | 0.178 |
|  | ***n (%)*** | ***n (%)*** | ***Mann-Whitney p-value*** |
| **Sex** |  |  |  |
| Male | 981 (55.0%) | 799 (55.1%) | 0.965 |
| Female | 803 (45.0%) | 652 (44.9%) |  |
| **Type of onset** |  |  |  |
| Bulbar onset | 598 (33.5%) | 489 (33.7%) | 0.808 |
| Spinal onset | 1186 (65.5%) | 962 (66.3%) |  |

**eTABLE 2. Summary of complete blood count (CBC) values and derived inflammatory markers.**

|  | **Total** (median, (IQR)) | **Sex** (median, (IQR)) | | **Site of onset** (median, (IQR)) | |
| --- | --- | --- | --- | --- | --- |
|  | (n=1451) | **Male (n=799)** | **Female (n=652)** | **Bulbar (n=489)** | **Spinal (n=962)** |
| **WBC** | 6.27 (5.24-7.60) | 6.41 (5.39-7.72) | 6.14 (5.05-7.43) | 6.04 (5.11-7.54) | 6.4 (5.31-7.63) |
| **Neutrophils** | 3.77 (2.92-4.88) | 3.85 (3.10-4.97) | 3.64 (2.78-4.72) | 3.63 (2.80-4.81) | 3.82 (2.96-4.88) |
| **Lymphocytes** | 1.72 (1.36-2.17) | 1.71 (1.37-2.13) | 1.74 (1.36-2.19) | 1.67 (1.34-2.15) | 1.74 (1.37-2.18) |
| **Monocytes** | 0.48 (0.38-0.61) | 0.5 (0.40-0.65) | 0.45 (0.34-0.57) | 0.47 (0.37-0.60) | 0.49 (0.38-0.62) |
| **Platelets** | 229.5 (193.0-274.0) | 216 (180.0-259.0) | 246 (209.0-292.0) | 220 (184.0-272.0) | 233 (196.0-275.0) |
| **NLR** | 2.1 (1.6-3.1) | 2.2 (1.7-3.1) | 2.0 (1.48-2.97) | 2.1 (1.54-3.03) | 2.15 (1.60-3.08) |
| **PLR** | 134.6 (101.2-176.5) | 126.0 (97.7-164.9) | 142.5 (109.2-189.6) | 134.9 (103.6-177.1) | 134.39 (100.7-176.0) |
| **SII** | 495.37 (340.3-746.2) | 485.75 (338.6-720.8) | 510.9 (342.8-7567.0) | 479.6 (324.4-723.5) | 506.7 (343.9-749.0) |
| **LMR** | 3.58 (2.7-4.79) | 3.89 (2.96-5.38) | 3.45 (2.67-4.69) | 3.61 (2.73-4.79) | 3.56 (2.69-4.8) |

**eTABLE 3A. Influence of smoking status on CBC data and inflammatory markers.**

|  | **Active smoking** | | |
| --- | --- | --- | --- |
| *Predictors* | *OR* | *CI* | *p* |
| Neutrophil | 1.03 | 1.01 – 1.06 | **0.01** |
| Monocyte | 1 | 0.98 – 1.03 | 0.697 |
| Platelet | 1.01 | 0.98 – 1.03 | 0.65 |
| NLR | 0.99 | 0.97 – 1.01 | 0.415 |
| PLR | 0.99 | 0.97 – 1.01 | 0.233 |
| SII | 0.99 | 0.96 – 1.01 | 0.298 |
| Lymphocyte | 1.02 | 1.00 – 1.04 | 0.06 |
| LMR | 1 | 0.98 – 1.02 | 0.813 |

**eTABLE 3B. Influence of bulbar symptoms on CBC data and inflammatory markers.**

|  | **Bulbar symptoms** | | |
| --- | --- | --- | --- |
| *Predictors* | *OR* | *CI* | *p* |
| Neutrophil | 1.01 | 0.98 – 1.04 | 0.688 |
| Monocyte | 0.99 | 0.92 – 1.07 | 0.842 |
| Platelet | 1.01 | 0.98 – 1.04 | 0.54 |
| NLR | 1 | 0.97 – 1.03 | 0.935 |
| PLR | 0.99 | 0.90 – 1.10 | 0.902 |
| SII | 1.01 | 0.97 – 1.04 | 0.742 |
| Lymphocyte | 1 | 0.97 – 1.02 | 0.8 |
| LMR | 1 | 0.97 – 1.03 | 0.932 |

**eTABLE 4. Summary of multivariate analysis of immune and adaptive inflammation markers in the whole cohort** (OR odds ratio, HR hazard ratio, 97.5% CI 97.55% confidence interval, NLR neutrophil-to-lymphocyte ratio, PLR platelet-to-lymphocyte ratio, SII systemic inflammatory index, LMR lymphocyte-to-monocyte ratio)

|  | **FVC < 75%** | | |
| --- | --- | --- | --- |
| *Predictors* | *OR* | *97.5% CI* | *P* |
| Neutrophil | 1.06 | 1.03 – 1.09 | **<0.001** |
| Monocyte | 1.01 | 0.93 – 1.09 | 0.834 |
| Platelet | 1.02 | 0.99 – 1.06 | 0.216 |
| NLR | 1.07 | 1.03 – 1.10 | **<0.001** |
| PLR | 1.15 | 1.04 – 1.28 | **0.007** |
| SII | 1.09 | 1.04 – 1.13 | **<0.001** |
| Lymphocyte | 0.97 | 0.95 – 1.00 | **0.03** |
| LMR | 1 | 0.97 – 1.02 | 0.899 |
|  | **ALSFRS-R PROGRESSION RATE** | | |
| *Predictors* | *β* | *97.5% CI* | *p* |
| Neutrophil | 1.03 | 1.01 – 1.05 | **0.001** |
| Monocyte | 1.05 | 1.00 – 1.10 | **0.041** |
| Platelet | 1 | 0.98 – 1.02 | 0.801 |
| NLR | 1.03 | 1.01 – 1.05 | **0.008** |
| PLR | 1.05 | 0.99 – 1.11 | 0.136 |
| SII | 1.03 | 1.01 – 1.05 | **0.006** |
| Lymphocyte | 0.99 | 0.98 – 1.01 | 0.413 |
| LMR | 0.99 | 0.98 – 1.01 | 0.263 |
|  | **COGNITIVE IMPAIRMENT** | | |
| *Predictors* | *OR* | *97.5% CI* | *p* |
| Neutrophil | 1.01 | 0.95 – 1.07 | 0.724 |
| Monocyte | 0.84 | 0.73 – 0.97 | **0.017** |
| Platelet | 1 | 0.94 – 1.07 | 0.975 |
| NLR | 1.05 | 0.98 – 1.12 | 0.171 |
| PLR | 1.16 | 0.94 – 1.44 | 0.167 |
| SII | 1.07 | 0.99 – 1.16 | 0.093 |
| Lymphocyte | 0.97 | 0.93 – 1.02 | 0.225 |
| LMR | 1.03 | 0.99 – 1.08 | 0.166 |
|  | **FTD** | | |
| *Predictors* | *OR* | *97.5% CI* | *P* |
| Neutrophil | 1 | 0.97 – 1.03 | 0.928 |
| Monocyte | 0.92 | 0.86 – 0.99 | **0.036** |
| Platelet | 1 | 0.97 – 1.03 | 0.985 |
| NLR | 1.03 | 0.99 – 1.06 | 0.134 |
| PLR | 1.11 | 0.99 – 1.24 | 0.068 |
| SII | 1.03 | 0.99 – 1.07 | 0.138 |
| Lymphocyte | 0.99 | 0.97 – 1.01 | 0.259 |
| LMR | 1.02 | 0.99 – 1.04 | 0.138 |
|  | **SURVIVAL** | | |
| *Predictors* | *HR* | *97.5% CI* | *P* |
| Neutrophil | 1.11 | 1.05 – 1.17 | **<0.001** |
| Monocyte | 1.02 | 0.97 – 1.07 | 0.366 |
| Platelet | 0.99 | 0.93 – 1.04 | 0.616 |
| NLR | 1.13 | 1.07 – 1.18 | **<0.001** |
| PLR | 1.01 | 0.96 – 1.07 | 0.67 |
| SII | 1.12 | 1.06 – 1.17 | **<0.001** |
| Lymphocyte | 0.96 | 0.88 – 1.05 | 0.419 |
| LMR | 0.95 | 0.89 – 1.02 | 0.14 |

**eTABLE 5. Summary of multivariate analysis of immune and adaptive inflammation markers according to sex** (OR odds ratio, HR hazard ratio, 97.5% CI 97.55% confidence interval, NLR neutrophil-to-lymphocyte ratio, PLR platelet-to-lymphocyte ratio, SII systemic inflammatory index, LMR lymphocyte-to-monocyte ratio)

|  |  | **FVC < 75%** | | |  |  |
| --- | --- | --- | --- | --- | --- | --- |
|  | **Female** | | | **Male** | | |
| *Predictors* | *OR* | *97.5% CI* | *p* | *Estimates* | *97.5% CI* | *p* |
| Neutrophil | 1.05 | 1.00 – 1.10 | **0.036** | 1.07 | 1.02 – 1.12 | **0.005** |
| Monocyte | 0.97 | 0.85 – 1.11 | 0.641 | 1.03 | 0.93 – 1.13 | 0.573 |
| Platelet | 1.03 | 0.98 – 1.09 | 0.217 | 1.01 | 0.97 – 1.06 | 0.594 |
| NLR | 1.08 | 1.03 – 1.13 | **0.001** | 1.05 | 1.00 – 1.10 | **0.033** |
| PLR | 1.35 | 1.15 – 1.59 | **<0.001** | 1.05 | 0.92 – 1.19 | 0.488 |
| SII | 1.14 | 1.07 – 1.21 | **<0.001** | 1.06 | 1.00 – 1.12 | **0.039** |
| Lymphocyte | 0.91 | 0.83 – 0.98 | **0.018** | 0.99 | 0.96 – 1.02 | 0.438 |
| LMR | 0.97 | 0.91 – 1.04 | 0.36 | 1 | 0.98 – 1.03 | 0.8 |
|  |  | **ALSFRS-R PROGRESSION RATE** | | |  |  |
|  | **Female** | | | **Male** | | |
| *Predictors* | *β* | *97.5% CI* | *p* | *Estimates* | *97.5% CI* | *p* |
| Neutrophil | 1.03 | 1.01 – 1.05 | **0.012** | 1.05 | 1.01 – 1.08 | **0.004** |
| Monocyte | 1.09 | 1.02 – 1.16 | **0.009** | 1.06 | 0.99 – 1.12 | 0.083 |
| Platelet | 0.99 | 0.97 – 1.02 | 0.688 | 1 | 0.97 – 1.03 | 0.955 |
| NLR | 1.03 | 1.00 – 1.05 | **0.02** | 1.04 | 1.01 – 1.08 | **0.011** |
| PLR | 1 | 0.97 – 1.03 | 0.851 | 1.06 | 0.98 – 1.15 | 0.173 |
| SII | 1.04 | 1.01 – 1.06 | **0.007** | 1.03 | 1.00 – 1.07 | **0.039** |
| Lymphocyte | 0.97 | 0.93 – 1.01 | 0.129 | 0.99 | 0.97 – 1.01 | 0.415 |
| LMR | 0.97 | 0.94 – 1.00 | **0.049** | 0.99 | 0.97 – 1.01 | 0.413 |
|  |  | **COGNITIVE IMPAIRMENT** | | |  |  |
|  | **Female** | | | **Male** | | |
| *Predictors* | *OR* | *97.5% CI* | *p* | *Estimates* | *97.5% CI* | *p* |
| Neutrophil | 0.97 | 0.90 – 1.05 | 0.461 | 1.06 | 0.97 – 1.16 | 0.201 |
| Monocyte | 0.79 | 0.63 – 1.00 | **0.046** | 0.88 | 0.73 – 1.05 | 0.146 |
| Platelet | 1.01 | 0.92 – 1.12 | 0.778 | 0.99 | 0.91 – 1.07 | 0.754 |
| NLR | 1.03 | 0.94 – 1.12 | 0.591 | 1.07 | 0.97 – 1.19 | 0.168 |
| PLR | 1.28 | 0.95 – 1.73 | 0.106 | 1.02 | 0.76 – 1.38 | 0.885 |
| SII | 1.05 | 0.95 – 1.17 | 0.345 | 1.08 | 0.96 – 1.22 | 0.201 |
| Lymphocyte | 0.89 | 0.77 – 1.02 | 0.096 | 0.98 | 0.94 – 1.03 | 0.432 |
| LMR | 1.07 | 0.95 – 1.20 | 0.24 | 1.02 | 0.98 – 1.07 | 0.312 |
|  |  | **FTD** | | |  |  |
|  | **Female** | | | **Male** | | |
| *Predictors* | *OR* | *97.5% CI* | *p* | *Estimates* | *97.5% CI* | *p* |
| Neutrophil | 0.98 | 0.94 – 1.02 | 0.291 | 1.03 | 0.98 – 1.08 | 0.209 |
| Monocyte | 0.87 | 0.77 – 0.98 | **0.023** | 0.96 | 0.88 – 1.05 | 0.388 |
| Platelet | 0.99 | 0.94 – 1.05 | 0.783 | 1 | 0.96 – 1.04 | 0.981 |
| NLR | 1.02 | 0.97 – 1.07 | 0.502 | 1.04 | 0.99 – 1.09 | 0.145 |
| PLR | 1.19 | 1.01 – 1.40 | **0.036** | 1.02 | 0.88 – 1.19 | 0.774 |
| SII | 1.02 | 0.97 – 1.08 | 0.451 | 1.04 | 0.98 – 1.10 | 0.245 |
| Lymphocyte | 0.91 | 0.85 – 0.99 | **0.022** | 0.99 | 0.97 – 1.02 | 0.599 |
| LMR | 1.02 | 0.96 – 1.08 | 0.548 | 1.02 | 0.99 – 1.04 | 0.184 |
|  |  | **SURVIVAL** | | |  |  |
|  | **Female** | | | **Male** | | |
| *Predictors* | *HR* | *97.5% CI* | *p* | *Estimates* | *97.5% CI* | *p* |
| Neutrophil | 1.09 | 1.01 – 1.19 | **0.03** | 1.07 | 0.97 – 1.17 | 0.165 |
| Monocyte | 1.17 | 0.92 – 1.49 | 0.197 | 0.98 | 0.79 – 1.20 | 0.815 |
| Platelet | 1.08 | 0.97 – 1.20 | 0.154 | 0.95 | 0.86 – 1.03 | 0.22 |
| NLR | 1.15 | 1.07 – 1.25 | **<0.001** | 1.11 | 1.01 – 1.22 | **0.028** |
| PLR | 1.04 | 0.95 – 1.13 | 0.392 | 1.08 | 0.84 – 1.39 | 0.558 |
| SII | 1.3 | 1.18 – 1.44 | **<0.001** | 1.07 | 0.97 – 1.18 | 0.156 |
| Lymphocyte | 0.84 | 0.70 – 1.00 | **0.045** | 1 | 0.91 – 1.09 | 0.991 |
| LMR | 0.83 | 0.72 – 0.96 | **0.013** | 1.02 | 0.95 – 1.08 | 0.639 |

**eTABLE 6. Summary of multivariate analysis of immune and adaptive inflammation markers according to age group (< 60 years, 60-70 years, > 70 years)** (OR odds ratio, HR hazard ratio, 97.5% CI 97.55% confidence interval, NLR neutrophil-to-lymphocyte ratio, PLR platelet-to-lymphocyte ratio, SII systemic inflammatory index, LMR lymphocyte-to-monocyte ratio)

|  | **FVC < 75%** | | | | | | | | |
| --- | --- | --- | --- | --- | --- | --- | --- | --- | --- |
|  | **< 60 years** | | | **60-70 years** | | | **> 70 years** | | |
| *Predictors* | *OR* | *97.5% CI* | *P* | *Estimates* | *97.5% CI* | *p* | *Estimates* | *97.5% CI* | *p* |
| Neutrophil | 1.06 | 1.00 – 1.12 | 0.066 | 1.05 | 1.01 – 1.10 | **0.026** | 1.08 | 1.00 – 1.18 | 0.052 |
| Monocyte | 1.01 | 0.89 – 1.15 | 0.877 | 0.97 | 0.86 – 1.09 | 0.619 | 1.12 | 0.93 – 1.34 | 0.236 |
| Platelet | 1.02 | 0.96 – 1.08 | 0.487 | 1.03 | 0.98 – 1.08 | 0.269 | 1.02 | 0.94 – 1.11 | 0.649 |
| NLR | 1.06 | 0.98 – 1.14 | 0.125 | 1.06 | 1.01 – 1.11 | **0.021** | 1.08 | 1.01 – 1.16 | **0.019** |
| PLR | 1.15 | 0.95 – 1.39 | 0.165 | 1.12 | 0.97 – 1.29 | 0.117 | 1.29 | 1.01 – 1.64 | **0.043** |
| SII | 1.09 | 1.00 – 1.17 | **0.038** | 1.07 | 1.01 – 1.13 | **0.013** | 1.15 | 1.04 – 1.27 | **0.005** |
| Lymphocyte | 0.97 | 0.89 – 1.05 | 0.395 | 0.99 | 0.96 – 1.02 | 0.465 | 0.89 | 0.78 – 1.01 | 0.065 |
| LMR | 1.01 | 0.97 – 1.06 | 0.522 | 1 | 0.97 – 1.04 | 0.903 | 0.94 | 0.85 – 1.03 | 0.19 |
|  | **ALSFRS-R PROGRESSION RATE** | | | | | | | | |
|  | **< 60 years** | | | **60-70 years** | | | **> 70 years** | | |
| *Predictors* | *β* | *97.5% CI* | *p* | *Estimates* | *97.5% CI* | *p* | *Estimates* | *97.5% CI* | *p* |
| Neutrophil | 1.01 | 0.96 – 1.05 | 0.774 | 1.05 | 1.02 – 1.07 | **<0.001** | 1.03 | 0.98 – 1.07 | 0.228 |
| Monocyte | 1.01 | 0.92 – 1.10 | 0.886 | 1.09 | 1.03 – 1.16 | **0.005** | 1.03 | 0.94 – 1.13 | 0.507 |
| Platelet | 0.97 | 0.93 – 1.01 | 0.089 | 1.02 | 0.99 – 1.05 | 0.135 | 1.01 | 0.97 – 1.05 | 0.62 |
| NLR | 1.02 | 0.97 – 1.08 | 0.488 | 1.04 | 1.02 – 1.07 | **<0.001** | 1 | 0.96 – 1.04 | 0.977 |
| PLR | 1.01 | 0.88 – 1.15 | 0.933 | 1.08 | 1.01 – 1.16 | **0.026** | 1.01 | 0.89 – 1.14 | 0.905 |
| SII | 1.01 | 0.95 – 1.06 | 0.833 | 1.05 | 1.02 – 1.07 | **<0.001** | 1.01 | 0.97 – 1.06 | 0.529 |
| Lymphocyte | 0.96 | 0.91 – 1.02 | 0.196 | 0.99 | 0.98 – 1.01 | 0.308 | 1.07 | 0.99 – 1.15 | 0.102 |
| LMR | 0.99 | 0.96 – 1.03 | 0.679 | 0.99 | 0.97 – 1.00 | 0.139 | 1.02 | 0.96 – 1.08 | 0.502 |
|  | **COGNITIVE IMPAIRMENT** | | | | | | | | |
|  | **< 60 years** | | | **60-70 years** | | | **> 70 years** | | |
| *Predictors* | *OR* | *CI* | *p* | *Estimates* | *97.5% CI* | *p* | *Estimates* | *97.5% CI* | *p* |
| Neutrophil | 1.05 | 0.94 – 1.18 | 0.366 | 1 | 0.93 – 1.08 | 0.991 | 1.03 | 0.91 – 1.17 | 0.646 |
| Monocyte | 0.79 | 0.63 – 0.99 | **0.041** | 0.84 | 0.67 – 1.04 | 0.105 | 0.85 | 0.61 – 1.20 | 0.35 |
| Platelet | 1.03 | 0.92 – 1.16 | 0.584 | 0.99 | 0.90 – 1.08 | 0.804 | 1 | 0.85 – 1.18 | 0.98 |
| NLR | 1.05 | 0.96 – 1.15 | 0.316 | 1.01 | 0.93 – 1.10 | 0.756 | 1.21 | 0.98 – 1.50 | 0.073 |
| PLR | 1.11 | 0.76 – 1.62 | 0.577 | 1.12 | 0.83 – 1.53 | 0.455 | 1.49 | 0.92 – 2.43 | 0.104 |
| SII | 1.13 | 0.96 – 1.33 | 0.143 | 1.04 | 0.95 – 1.15 | 0.394 | 1.16 | 0.96 – 1.41 | 0.122 |
| Lymphocyte | 0.99 | 0.86 – 1.14 | 0.874 | 0.98 | 0.93 – 1.03 | 0.39 | 0.7 | 0.54 – 0.90 | **0.006** |
| LMR | 1.11 | 1.03 – 1.19 | **0.006** | 1.01 | 0.96 – 1.07 | 0.657 | 0.85 | 0.66 – 1.10 | 0.205 |
|  | **FTD** | | | | | | | | |
|  | **< 60 years** | | | **60-70 years** | | | **> 70 years** | | |
| *Predictors* | *OR* | *97.5% CI* | *p* | *Estimates* | *97.5% CI* | *p* | *Estimates* | *97.5% CI* | *p* |
| Neutrophil | 1.03 | 0.97 – 1.09 | 0.366 | 0.99 | 0.96 – 1.03 | 0.73 | 1.03 | 0.96 – 1.11 | 0.377 |
| Monocyte | 0.9 | 0.81 – 1.01 | 0.062 | 0.93 | 0.83 – 1.04 | 0.219 | 0.91 | 0.75 – 1.11 | 0.357 |
| Platelet | 1.03 | 0.98 – 1.09 | 0.253 | 0.99 | 0.94 – 1.04 | 0.637 | 1 | 0.91 – 1.10 | 0.947 |
| NLR | 1.01 | 0.97 – 1.06 | 0.561 | 1.01 | 0.97 – 1.06 | 0.6 | 1.15 | 1.02 – 1.30 | **0.021** |
| PLR | 1.07 | 0.89 – 1.29 | 0.464 | 1.1 | 0.94 – 1.28 | 0.256 | 1.31 | 0.99 – 1.74 | 0.057 |
| SII | 1.06 | 0.98 – 1.15 | 0.163 | 1.01 | 0.96 – 1.07 | 0.645 | 1.13 | 1.01 – 1.26 | **0.03** |
| Lymphocyte | 0.99 | 0.93 – 1.06 | 0.801 | 0.99 | 0.97 – 1.02 | 0.536 | 0.82 | 0.71 – 0.94 | **0.006** |
| LMR | 1.05 | 1.01 – 1.09 | **0.006** | 1.01 | 0.98 – 1.04 | 0.515 | 0.93 | 0.80 – 1.07 | 0.302 |
|  | **SURVIVAL** | | | | | | | | |
|  | **< 60 years** | | | **60-70 years** | | | **> 70 years** | | |
| *Predictors* | *HR* | *97.5% CI* | *p* | *Estimates* | *97.5% CI* | *p* | *Estimates* | *97.5% CI* | *P* |
| Neutrophil | 1.02 | 0.91 – 1.15 | 0.678 | 1.08 | 1.00 – 1.17 | **0.04** | 1.16 | 1.05 – 1.29 | **0.005** |
| Monocyte | 0.94 | 0.74 – 1.20 | 0.628 | 1.02 | 0.93 – 1.12 | 0.657 | 1 | 0.94 – 1.06 | 0.984 |
| Platelet | 1.01 | 0.91 – 1.13 | 0.852 | 0.98 | 0.90 – 1.08 | 0.735 | 1.01 | 0.91 – 1.12 | 0.842 |
| NLR | 1.07 | 0.96 – 1.21 | 0.234 | 1.13 | 1.05 – 1.21 | **0.001** | 1.1 | 1.00 – 1.21 | **0.047** |
| PLR | 1.03 | 0.93 – 1.15 | 0.563 | 1 | 0.92 – 1.07 | 0.914 | 1.21 | 0.86 – 1.70 | 0.273 |
| SII | 1.1 | 0.98 – 1.22 | 0.094 | 1.09 | 1.01 – 1.17 | **0.022** | 1.19 | 1.06 – 1.32 | **0.002** |
| Lymphocyte | 0.92 | 0.77 – 1.09 | 0.331 | 1.02 | 0.95 – 1.10 | 0.581 | 0.99 | 0.79 – 1.24 | 0.936 |
| LMR | 1 | 0.91 – 1.10 | 0.966 | 0.97 | 0.88 – 1.07 | 0.545 | 1.02 | 0.85 – 1.24 | 0.804 |
